# Quantifying the relationship between SARS-CoV-2 viral load and infectiousness

**DOI:** 10.1101/2021.05.07.21256341

**Authors:** Aurélien Marc, Marion Kerioui, François Blanquart, Julie Bertrand, Oriol Mitjà, Marc Corbacho-Monné, Michael Marks, Jérémie Guedj

**Affiliations:** Université de Paris, IAME, INSERM, F-75018 Paris, France; Lihir Medical Centre, International SOS, Lihir Island, Papua New Guinea; Fight AIDS and Infectious Diseases Foundation, Hospital Universitari Germans Trias i Pujol, Badalona, Spain; Hospital Universitari Parc Taulí, I3PT, Sabadell, Spain; Facultat de Medicina–Universitat de Barcelona, Barcelona, Spain; London School of Hygiene and Tropical Medicine, London, United Kingdom; Hospital for Tropical Diseases, London, United Kingdom

## Abstract

The relationship between SARS-CoV-2 viral load and infectiousness is not known. Using data from a prospective cohort of index cases and high-risk contact, we reconstructed by modelling the viral load at the time of contact and the probability of infection. The effect of viral load was particularly large in household contacts, with a transmission probability that increased to as much as 37% when the viral load was greater than 10 log_10_ copies per mL. The transmission probability peaked at symptom onset in most individuals, with a median probability of transmission of 15%, that hindered large individual variations (IQR: [8, 37]). The model also projects the effects of variants on disease transmission. Based on the current knowledge that viral load is increased by 2 to 4-fold on average, we estimate that infection with B1.1.7 virus could lead to an increase in the probability of transmission by 8 to 17%.

## Introduction

After more than 16 months of an unprecedented pandemic, some key aspects of disease transmission remain poorly understood. While respiratory droplets and aerosols have been rapidly demonstrated to be a major route of transmission of SARS-CoV-2^1^, the role of the viral load as a driver of infectiousness has been suspected but not formally established. This lack of evidence is due to the fact that documented high-risk contacts occur mostly before the index has been diagnosed, with no information on the viral load level at the time of the contact. The relationship between viral load and infectiousness determines the timing of transmission, part of the inter-individual heterogeneity in transmission, and the impact of interventions (contact / case isolation, vaccination) on transmission. In the context of variants of concern^2,3^, that are likely associated with larger viral loads, it becomes even more critical to delineate the contribution of viral shedding from other suspected factors associated with an increased transmission. Further, as antiviral and vaccine strategies are being implemented, that directly reduce the amount of viral shedding^4^, it is essential to understand how they may contribute to a reduction in SARS-CoV-2 transmission.

One of the most exhaustive clinical study to address the question of viral load and infectiousness has been obtained through individuals included in a randomised controlled trial done in March-April 2020 in Spain, that aimed to assess the efficacy of hydroxychloroquine on SARS-CoV-2 transmission^5^. Overall, 282 index and their 753 high-risk contacts were frequently monitored to assess their virological and clinical evolution, as well as possible infection for the contact. Interestingly, a clear association was found between the probability of being infected after a high-risk contact and the viral load measured at the time of diagnosis^5^. This suggests that viral load is associated with transmission; however, it does not quantify the role of viral load in disease transmission, as the viral load at the exact time of the contact remains unknown and may greatly differ from that measured, several days later, at the time of diagnosis.

In order to study in detail the role of viral load on the probability of transmission, we reanalysed these data by using a within-host model of viral dynamics^6,7^ to reconstruct the viral load levels of the index cases at the time of contact, and to infer the relationship between viral load and the probability of transmission after a high-risk contact. Further, we used the model to predict the effects of changes in viral load levels on the probability of transmission, representing the effects of infection with a variant of concern or infection in an individual in which vaccine would confer a partial protection against viral replication.

## Results

### Baseline characteristics

A total of 257 index cases and their 574 high-risk contacts (simply called contacts in the following) were included in this analysis (Supplementary Figure 1). A high risk contact was defined as a contact of >15 min within 2 meters of distance from a symptomatic case^8^.

The majority of index cases were female (72%) with a median age of 42 (interquartile range, IQR: [31, 52]). A total of 544 swab samples were performed at days 0, 3 and 7 days after study inclusion. Symptoms occurred at a median time of 4 days (IQR: [3, 5]) before the first swab sampling. The maximum median viral load obtained during follow was 8.4 log_10_ copies per mL (IQR: [6.9, 9.5]).

The majority of contacts were female (54%) with a median age of 41 (IQR: [28, 53]). The form of contacts was categorized as either household (60%) or non-household (40%).

The majority of contacts (65%) and of infection events (65%) occurred ±1 day from symptoms onset (Supplementary Figure 2).

Overall, 87 household contact led to an infection (proportion of transmission of 25.1%) and 29 non-household contacts led to an infection (proportion of transmission of 13%).

### Viral dynamic model

We used a target cell limited model to reconstruct the viral load kinetics of the index cases over time, assuming that the infection started 5 days before the onset of symptoms^6^. Although several models relating viral load to infectiousness were evaluated (see below), they all provided nearly identical fits to the viral load data predicted in the index cases (Figure 1).

**Figure 1:**
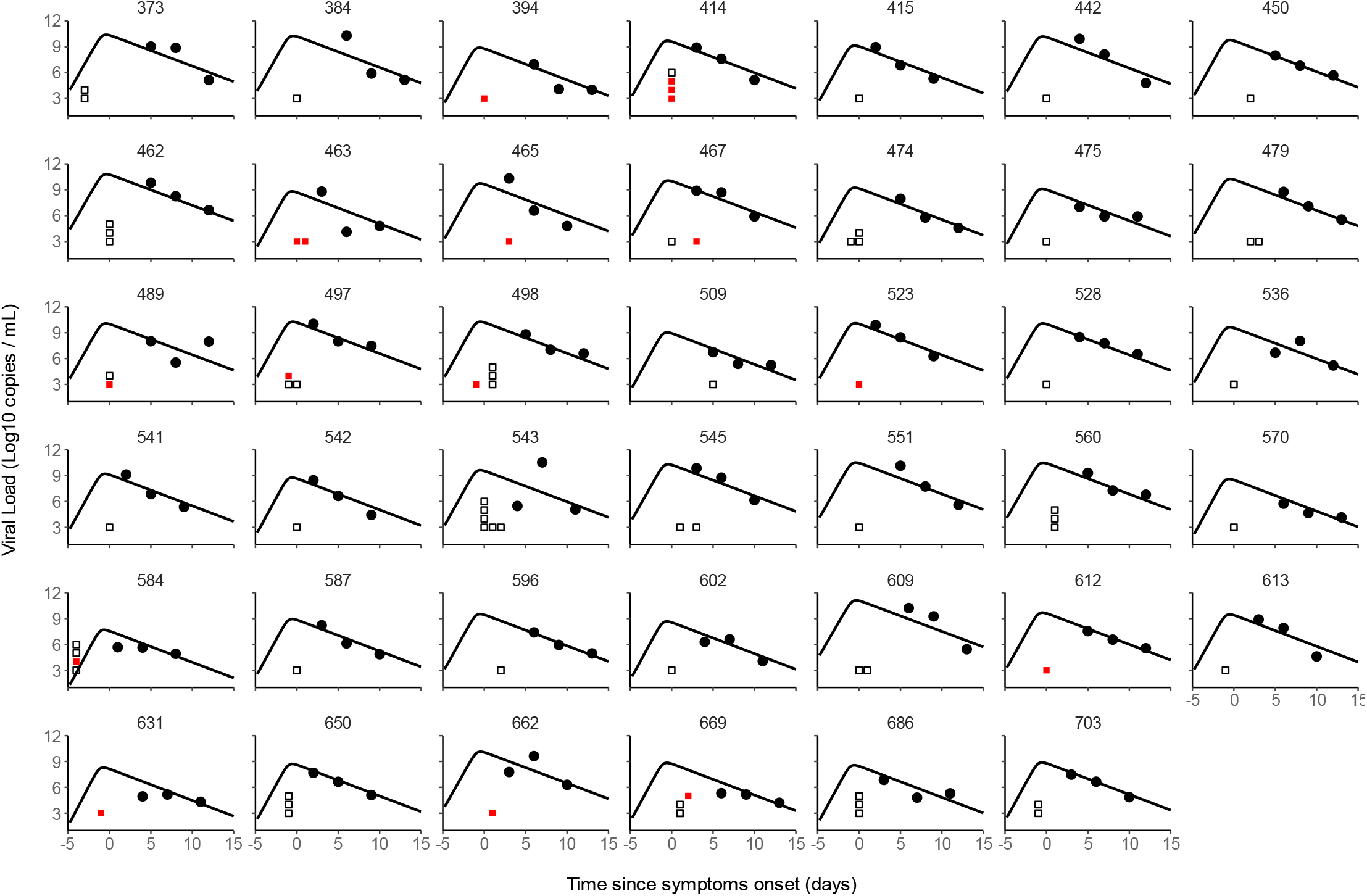
Individual prediction of viral kinetics for a subset of 41 index cases having 3 viral load measurements. Black dots represent the measured viral load. Empty squares represent contact without transmission. Red squares represent contact with transmission.

**Table 1 :** Parameters estimates of the three candidate models. R_0_, basic reproductive number; δ, loss rate of infected cells; p, rate of viral production; γ_1_ represents the effect of household contacts on the transmission probability; γ_0_ represents the effect of non-household contacts on the transmission probability. M1 is a model where the transmission probability does not depend on the viral load. M2 and M3 are models where the transmission probability depends on the viral load at the time of contact.

|  | Parameter estimates (RSE %) |  |  |  |  |  |
| --- | --- | --- | --- | --- | --- | --- |
|  | No effect of viral load (M1) |  | Logit-Linear (M2) |  | Log-linear Model (M3) |  |
|  | Fixed effect | Random effect SD | Fixed effect | Random effect SD | Fixed effect | Random effect SD |
| $R_0$ | 14.2 (80) | 0.27 (278) | 16.2 (17) | 0.23 (48) | 16.0 (43) | 0.28 (76) |
| $\delta$ ($d^{-1}$) | 0.85 (14) | 0.048 (77) | 0.83 (7) | 0.045(72) | 0.83 (5) | 0.042 (41) |
| $\overset{p}{(cells^{-1}.d^{-1})}$ | $3.3 \times 10^5$ (148) | 2.54 (28) | $4.1 \times 10^5$ (77) | 2.61 (8) | $3.7 \times 10^5$ (127) | 2.56 (10) |
| $\gamma_1$ | 1.24 (69) | 0.87(114) | 0.38 (20) | 0.98 (28) | 0.39 (7) | 0.6 (26) |
| $\gamma_2$ | 0.55 (121) | | 0.16 (43) | | 0.22 (13) | |
| $BIC$ | 2490 | | 2487 | | 2489 | |

In the final model (Model M2), the basic within-host reproductive number, *R*_0_, quantifying the number of cell infections that occur from a single infected cell at the beginning, was estimated to 16.2, the loss rate of productively infected cells, *δ*, to 0.83 d^−1^ (corresponding to a half-life of 20 hours) and viral production *p*, to 4.1 × 10^5^ cells^−1^.d^−1^ (Table 2).

When reconstructing the viral profiles, the model predicted that the median peak viral load coincided with symptoms onset, with a median peak value of 9.8 log_10_ copies per mL (IQR: [9.1, 10.4]).

### Modelling the probability of transmission after a high-risk contact

We tested several models of probability transmission (see Methods) and estimated the parameters of both viral dynamics and probability of transmission simultaneously. The two model assuming an effect of viral load on the probability of transmission (Model M2 and M3) provided an improvement in BIC as compared to the model M1, supporting an effect of viral load on the probability of infection. In this model, viral load was significantly associated with the probability of transmission after household contact (P<0.01, Wald test on γ_1_), however the effect was less clear after non-household transmission (P<0.05, Wald test on γ_2_).

Of note our models were associated with a large between-subject variability on the effect of the viral load, *β* (as measured by the standard deviation of the associated random effects, ω_*β*_) suggesting that several other factors are involved in transmission, besides viral load. To represent this variability, we sampled 1,000 individuals in the population distribution to get prediction intervals for the viral load and the probability of transmission.

Using the simulated individuals, we inferred the viral load levels and the mean probability of transmission for both household and non-household contacts, that we compared with the observed proportion of infection for different viral load level category (Figure 2). The mean probability of transmission increased from the fixed nominal value of 5% for viral load levels <10^6^ copies per mL, to as much as 37% and 17% for viral load ≥10^10^ copies per mL for household and non-household contacts respectively. This is in agreement with the value of 37% and 29% observed in the clinical study (Figure 2).

**Figure 2:**
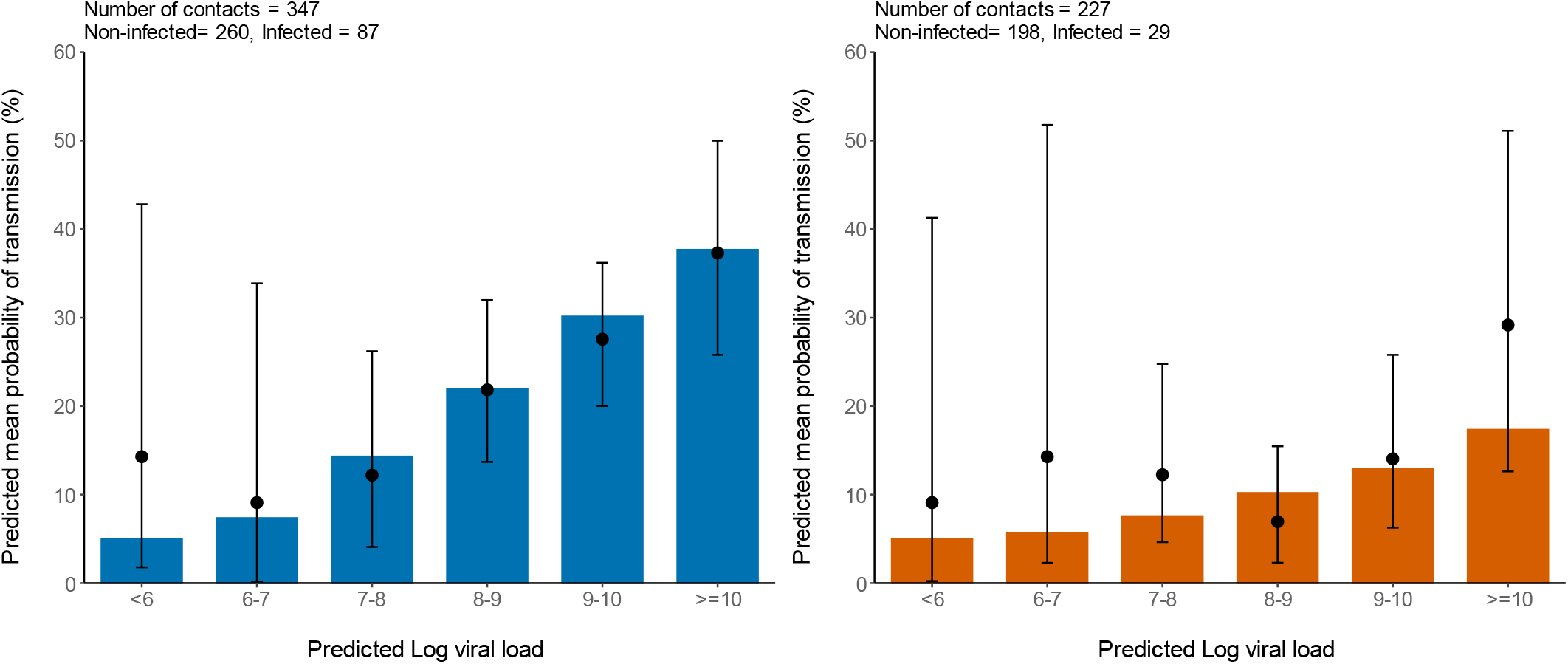
Mean probability of transmission according to different viral load levels of simulated individuals. The black dots represent the observed proportion of transmission. 95% confidence intervals are represented in black. Household contacts (Left). Non-household contacts (Right).

Over the time of infection, the probability of transmission peaked at the time of symptom onset, albeit with large inter-individual variabilities (Figure 3). In household contacts, the median peak of the probability of transmission was 15% (IQR: [8, 37]), but the mean value was much larger, to about 28%. The peak was much lower in non-household contacts, with a median value of 8% (IQR: [6, 13], and mean value of 13%. As a consequence of our assumption that the probability of transmission after a high-risk contact returned to baseline level when viral load dropped below the threshold of viral culture, i.e., 6 log_10_ copies per mL^6^, the window for infection was much shorter than the duration of viral shedding. The probability of transmission was above baseline during a median duration of 12 days (IQR: [10, 15]).

**Figure 3:**
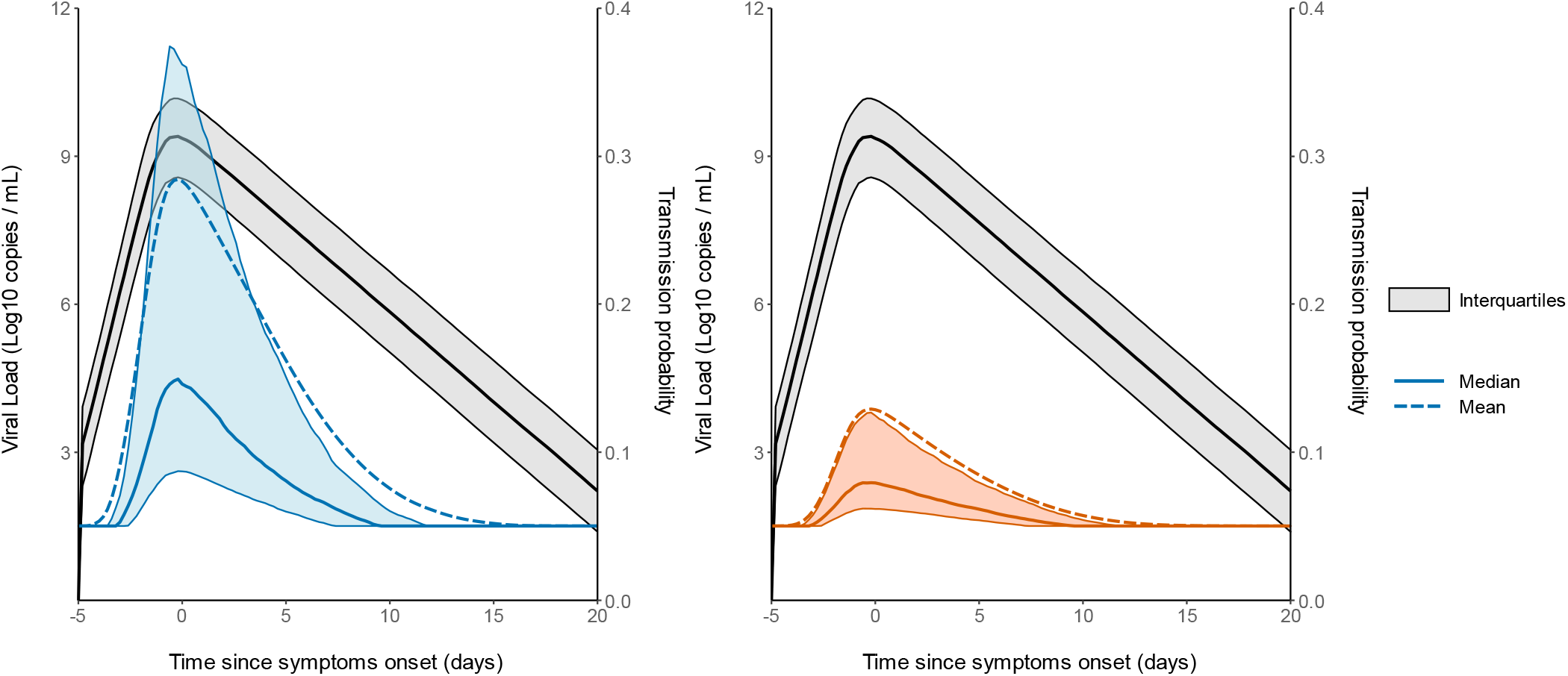
Prediction interval of the viral load and probability of transmission over time after a high-risk contact. The median and the mean are represented as a solid and dashed line respectively. (Black: Viral load. Probability of transmission for household contacts (Left). Probability of transmission for non-household contacts (Right). The shaded areas represent the interval between the first and third quartile. Realised on 1000 simulated individuals.

### Impact of changes in viral load levels on the probability of transmission

Finally, we used our model to characterize the effects of changes in viral load levels on the probability of transmission. For that purpose, we evaluated the impact of a change in the viral production rate, *p*, by a fold 2-100, which corresponds to an average increase in viral load of 1-7 cycle thresholds (Ct), at each time point.

To get a sense of the impact of these changes on infectiousness, we calculated the average probability of transmission after a high-risk contact in the overall population during the whole study period. We took into account the fact that contacts are not uniformly distributed, and we assumed a similar distribution of contacts as found in the original study for both household and non-household contacts (Supplementary Figure 2). For the baseline scenario using the estimated parameters in our population, the model accurately reproduced the observed transmission probability, at 25% and 11% for household and non-household contacts, respectively. With an increased value of viral production rate p by a factor 2, which corresponds to the viral load increase caused by B1.1.7 strain in large scale epidemiological studies^9–11^, the average probability of transmission would increase to 27% and 12% for household and non-household contacts respectively. With a 4-fold increase as suggested elsewhere^2^ the average probability of transmission would increase to 29% and 13%, respectively (Figure 4). The estimates for the P1 and B1.1.351 variants are much less well established, with values ranging from a 2-fold^2^ to a 10-fold increase^12^. Assuming an increase by 8-fold of the viral load, the average probability of transmission would increase to 31% and 14%, respectively, i.e., an increase of more than 25% from the baseline scenario (Figure 4 and Supplementary Table S2).

**Figure 4:**
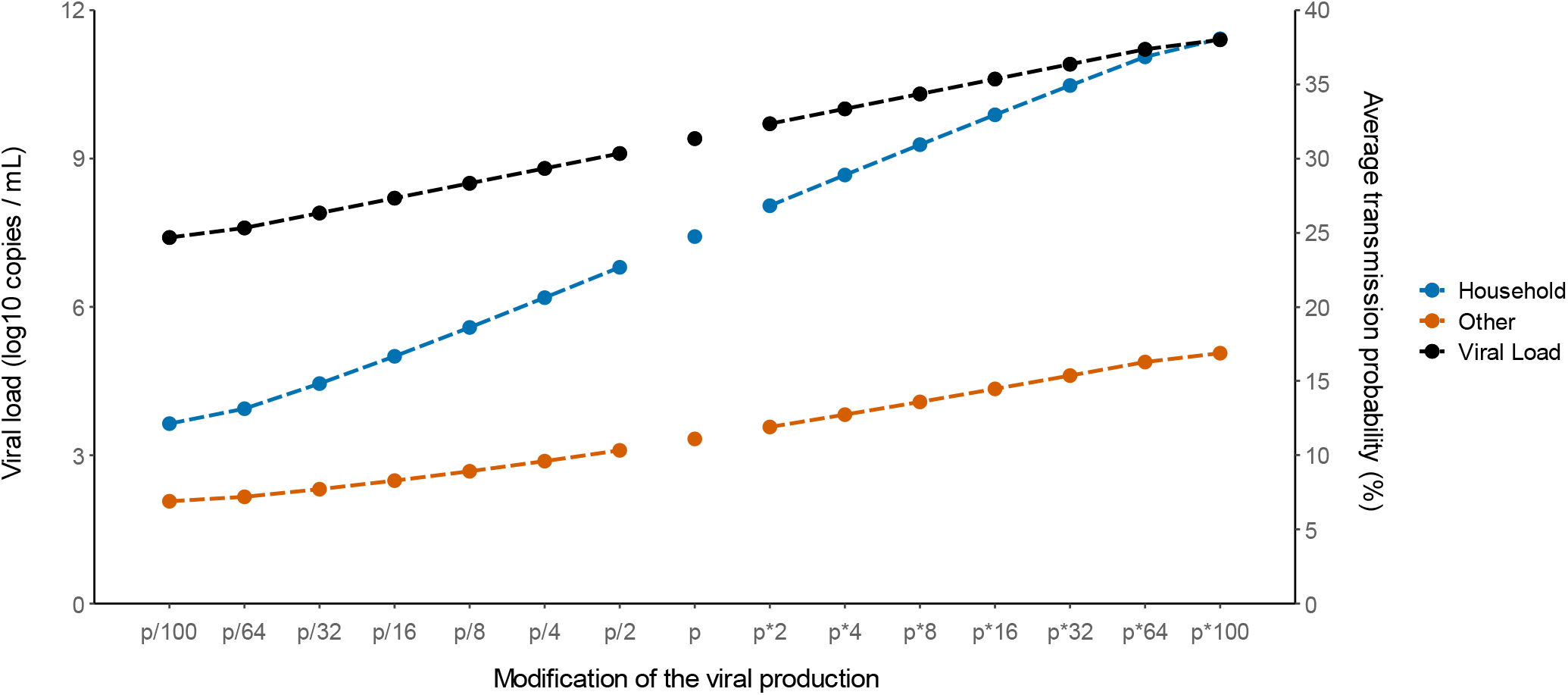
Impact of changes in the viral production rate on the average probability of transmission after household (blue) and non-household (blue) contacts. The black line represents the median peak viral load observed in each setting.

Conversely, we studied the effects of lower levels of viral load, as expected from a partial protection conferred by vaccination. Epidemiological studies in Israel reported a 3-5-fold lower viral load in infected vaccinated individuals as compared to unvaccinated individuals^4^. Assuming a reduction by a factor 4 of the viral production rate, p, would lead to an average probability of transmission of 21 and 10% for household and non-household contacts respectively. In another study relying on systematic repeated viral testing in both symptomatic and asymptomatic individuals, the effect of vaccine was much more dramatic, with a 100-fold reduction in viral load levels^13^. This would translate into an average transmission probability of 12 and 7%, which represents a decrease of 50% and 38% from the baseline scenario for household and non-household contacts respectively (Figure 4 and Supplementary Table S2).

In addition, we also tested the sensitivity of our results to the assumptions made for the effects of viral load. Exploring alternative models (M3) led to a largely similar result (Supplementary Table S2).

## Discussion

This is the first detailed description of the relationship between viral load and infectiousness. We here quantified the impact of viral load on infectiousness obtained on a highly detailed data obtained in a large epidemiological study^8^. The effect of viral load was particularly large in household contacts, with a mean transmission probability that increased to as much as 37% when the viral load was over 10 log_10_ copies per mL. Unlike what has been suggested until now by theoretical models^14,15^, the probability of transmission increased continuously with viral load and no saturation effects were visible at high viral loads (Supplementary Figure 3). However, and consistent with reports suggesting that the probability of transmission^16^ greatly vary between individuals, the effect of viral load was individual-dependent. For instance, at the peak of infectiousness, the median probability of transmission during household contact was 15%, but ranged from 5 to 100%.

The model also provided information on the effects of variants on disease transmission. We relied on results found in large-scale epidemiological data, that reported an average increase of the B1.1.7 virus by 1-2 Ct^2,9,10^. This can be reproduced in our model by assuming that viral production increases by a factor 2. Alternatively, as only the product p×T_0_ can be identified, this could also be due to B1.1.7 being able to infect twice as much target cells, as suggested by the fact that the N501Y substitution improved the affinity of the viral spike protein^3^. Regardless of the origin of this increased viral load, we estimated that an increase of viral load by a factor of 2 would increase the average transmission probability in household contacts from 25 to 27% (8% increase from baseline scenario). Assuming an increase by 4-fold of the viral load (2 Ct on average) would lead to a much more dramatic effect, increasing the average transmission probability in household contacts to 29% (17% increase from baseline scenario). Of note assuming a steeper effect of the viral load on the probability of transmission would lead to larger effect of variants (Supplementary Table S2).

Conversely, vaccination rollout is expected to confer a large level of protection, partly due to lower virus carriage in infected individuals. The exact magnitude of this decrease is difficult to quantify, and depends on the design of the studies, that include or not asymptomatic individuals. In fact preliminary reports have reported numbers going from a 5 to 100-fold reduction in viral load levels^13^. Whatever the exact value, it is clear that such reductions could be associated with large reductions in the probability of transmission.

Our study has some important limitations that need to be acknowledged. First, the reporting of high-risk contacts is partial and remains prone to various reporting biases. One of them is the fact that at the time where the study was conducted, there was no firm evidence of the role of pre-symptomatic transmission. This could explain why in our study a large number of high-risk household contact were reported to occur the day of symptom onset. It is also possible that several of the household contacts were not unique and occurred multiple times. Because we had no information on these contacts, we did not conduct specific analyses on repeated contacts, but it is something that future epidemiological studies will need to investigate. Another limitation is that we had no genomic data to ensure that infection observed in contact individuals results from an infection by the index case. In most infected contacts, we also did not have data on the time of symptom onset, which prevented us from detecting infections unlikely related to the contact identified in our study. However, the temporality of symptoms would not be sufficient to bring a decisive information on the infection event. Indeed, the study was conducted during the first epidemic wave in Spain, where most individuals, including in hospital settings, had not yet applied social distancing and masking, causing dozens of thousands of individuals infected every day. Both the possibility of repeated contacts in household and infection of contacts outside the identified contact network may have led us to overestimate the difference in the probability of transmission between household and non-household contacts. Specifically, infections outside of the identified probability contact would flatten the estimated relationship between viral load and transmission compared to the true relationship. It is also important to note that viral load data in index cases were collected on average 3-4 days after symptom onset, in the declining phase of viral load, several days after most of the contacts had occurred. To reconstruct the viral load profiles in absence of information, we assumed a fixed incubation period of 5 days with no variability^6,17^. As peak viral load could be related to symptom onset^5^, our approach of fixing a unique incubation period likely underestimates the variability of viral load dynamics in the pre-symptomatic phase. This may be partly compensated in our model by the large variability in the effect of viral load on transmission. Given the difficulty to access to very early virological data, this will remain a major limitation to this type of analysis. This can be partly studied in animal models^18^ but obviously at the cost of a very specific system with limited translation to human epidemiology.

To conclude our study quantifies the probability of infection according to viral load level after a high-risk contact. This relationship can be used to predict the effects of changes in virus paradigm, caused by the emergence of new variants and/or the rollout of vaccination. We estimate that 2-to 4-fold increase in viral load level observed with B1.1.7 virus could lead to an increase in the probability of transmission by 8 to 17% after a high-risk contact.

## Data Availability

The dataset used for this study is provided.

## Acknowledgements & Funding

The study has received financial support from the National Research Agency (ANR) through the ANR-Flash call for COVID-19 (Grant ANR-20-COVI-0018) and the Bill and Melinda Gates Foundation under Grant Agreement INV-017335. The original trial was funded by a crowdfunding campaign YoMeCorono (https://www.yomecorono.com/), and Generalitat de Catalunya with support for laboratory equipment from Foundation Dormeur. We thank Samuel Alizon, Xavier Duval and Xavier de Lamballerie for helpful discussions.

## Methods

### Data collection

Data used come from a cluster-randomized trial which included individuals with PCR-confirmed COVID-19 and their close contacts, and evaluated the efficacy of hydroxychloroquine as a pre- or post-exposure prophylaxis. The trial was conducted between March, 17 and April 28, 2020 in three out of nine health-care area in Catalonia, Spain. More details on the study protocol and main results can be found in the original publication^8^.

### Study participants

All index cases were aged 18 years or older with no hospitalisation, nasopharyngeal PCR positive results at baseline and mild symptoms onset within 5 days of inclusion. High-risk contacts were adults with a recent history of exposure and absence of COVID-19 like symptoms within the 7 days preceding enrolment. In the original publication, 282 index cases and the resulting 753 contacts were enrolled^5^; here we did not include 3 index individuals (and their corresponding 25 contacts) for which no viral load data was available, 8 index individuals (and their corresponding 19 contacts) for which no viral load was detected at any time point. Further, in 12 index cases (and their corresponding 127 contacts), no date of contact was available. Finally, we removed contacts occurring more than 5 days before symptoms onset, as they are unlikely to originate from the index case given the disease incubation time^6^. Thus, overall, our analysis was performed on 257 index and 574 contacts (see Supplementary Figure 1). In 12 index cases, the date of symptoms onset was not known and was imputed to 4 days before their first swab sampling, which corresponds to the median value observed in this population.

Type of contact was considered as household or non-household, which included Nursing home contacts, Health-care worker and other undefined contacts.

### Viral kinetic model

We used a target cell-limited model to reconstruct nasopharyngeal viral kinetics in index cases^6,19,20^. The model includes three populations of cells, namely Target cells (*T*), infected cells in their eclipse phase (*I*_1_) and productively infected cells (*I*_2_). Target cells (*T*) are infected at a constant rate *β* by infectious virus (*V*_*I*_). Infected cells enter an eclipse phase at a rate *k* before becoming productively infected cells (*I*_2_). We assumed productively infected cells have a constant loss rate *δ*. Virions are released from productively infected cells at a rate *p* and are loss at a rate *c*. A proportion *μ* of produced viruses are infectious (*V*_*I*_) and the remaining (1 − *μ*) are non-infectious viruses (*V*_*NI*_), both are cleared at a rate *c*. The model can be written as follows:

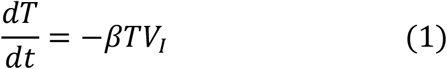

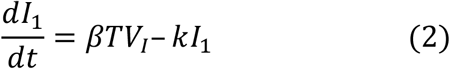

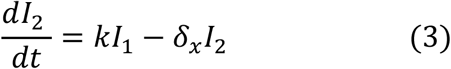

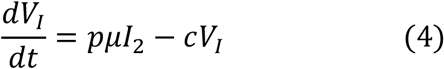

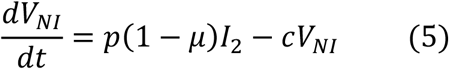

Based on this model, the basic reproduction number, *R*_0_, defined as the number of newly infected cells by one infected cell at the beginning of the infection^21^ is, 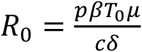. Given the absence of any antiviral effect of hydroxychloroquine against SARS-CoV-2^8,22,23^, we did not consider any effect of hydroxychloquine in the model.

### Assumptions on parameter values

Some parameters were fixed to ensure identifiability. The clearance rate *c* was fixed at 10 *d*^−1^ and the eclipse phase *k* to 4 *d*^−1^ based on previous work^6,7,24^. The proportion of infectious virus *μ* was assumed constant over time and was fixed to 10^−4^ as observed in animal model^24^. The initial number of target cells, *T*_0_, was calculated as follows: As there are 4 × 10^8^ epithelial cells in the upper respiratory tract (URT) and the volume of the URT is 30 *mL* we can calculate the concentration of epithelial cells in the 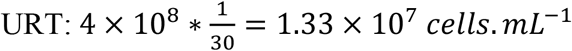. However, the ACE2 and TMPSS receptor needed for SARS-CoV-2 entry may be expressed by a small fraction of these cells. Hence, assuming the ACE2 receptor are expressed in 1%^25,26^ of these cells, we set T_0_= 1.33 × 10^5^ *cells. mL*^−1^. For each index case, the incubation period was fixed to 5 days as found in previous work, i.e., we assumed that the time of infection of index cases was exactly 5 days before symptom onset^6,17^. We assumed that at the moment of infection there was exactly one productively infected cell in the URT. Hence, 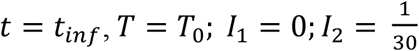; *V*_*I*_ = 0 *and V*_*NI*_ = 0.

### Statistical model for viral kinetics

Parameter estimations were performed using non-linear mixed-effect model. The structural model used to describe the observed log_10_ viral load is 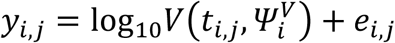 Where *y*_*i,j*_ is the j^*th*^ observation of index *i* at time *t*_*i,j*_ with *i* ∈ 1, ⃛, *N* and *j* ∈ 1, ⃛, *n*_*i*_ with *N* the number of index and *n*_*i*_ is the number of observations for index *i*. 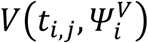 is the function describing the total viral load dynamics (*V*_*I*_(*t*_*i,j*_) + *V*_*NI*_(*t*_*i,j*_)) predicted by the model at time *t*_*i,j*_. The vector of viral kinetic parameters for index *i* is noted 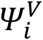 and *e*_*i,j*_ is the additive residual Gaussian error term of constant standard deviation *σ*. The vector of individual parameters depends on a fixed effects vector and on an individual random effects vector, which follows a normal centered distribution with a diagonal variance-covariance matrix *Ω*. To ensure positivity, the individual parameters follow a lognormal distribution.

### Probability of transmission

We noted 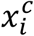 the *c*^*th*^ contact of index case *i* and *c* ∈ 1, ⃛, *C*_*i*_, with *C*_*i*_ the number of contacts of index *i*. The probability of transmission depends on the time of contact 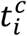, the nature of contact, namely household 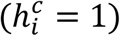 or not 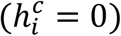, and the vector of individual parameters *Ψ*_*i*_, which contains the viral parameters 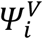 and transmission parameters *β*_*i*_ for index *i*. Five models of transmission were tested (M1-M5), described as follows:

#### Model M1

No effect of viral load

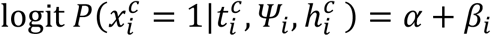

where: 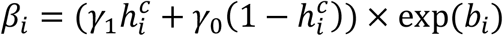 with γ_1_ (resp. γ_0_) is the effect of household contact (reps. non-household) on the probability of transmission, and *b*_*i*_ is an individual random effect assumed to follow a Gaussian distribution of variance 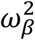. The baseline probability of transmission was fixed to 5% (*α*=-2.94).

#### Model M2

Logit-linear effect of viral load

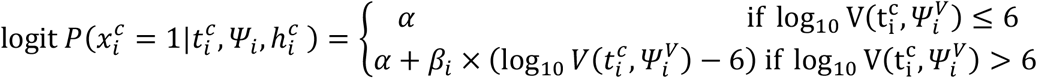

where: 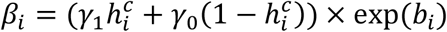 the effect of viral load on the probability of transmission in household contact (resp. non-household), and *b*_*i*_ a Gaussian individual random effect with variance 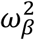. The baseline probability of transmission was fixed to 5% (α=-2.94) for viral load lower than 6 log_10_ copies per mL, which corresponds to the threshold for viral culture^6,15^.

#### Model M3

Log-linear effect of viral load

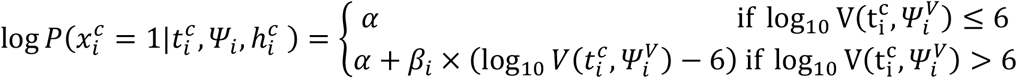

where: 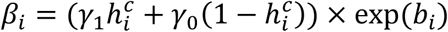 with γ_1_ (resp. γ_0_) the effect of viral load on the probability of transmission in household contact (resp. non-household), and *b*_*i*_ a Gaussian individual random effect with variance 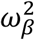. The baseline probability of transmission was fixed to 5% (α=-2.99) and the probability was bounded to 1.

#### Model M4

No effect of viral load with additive variability

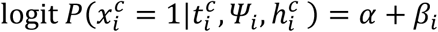

where: 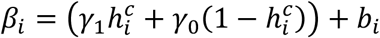 with γ_1_ (resp. γ_0_) is the effect of household contact (reps. non-household) on the probability of transmission, and *b*_*i*_ is an individual random effect assumed to follow a Gaussian distribution of variance 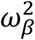. The baseline probability of transmission is fixed to 5% (*α*=-2.94).

#### Model M5

Logit-linear effect of the maximum predicted viral load

Finally, we tested the model M2 using the maximum of the predicted viral load.

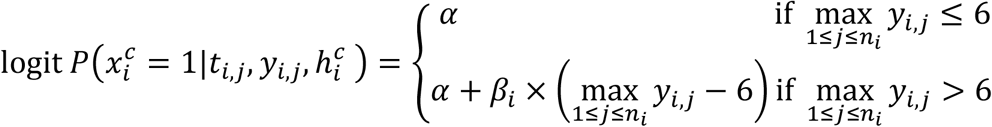

where: 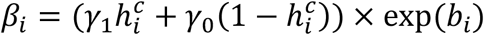 with γ_1_ (resp. γ_0_) the effect of viral load on the probability of transmission in household contact (resp. non-household), and *b*_*i*_ a Gaussian individual random effect with variance 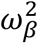. The baseline probability of transmission was fixed to 5% (α=-2.94) for viral load lower than 6 log_10_ copies per mL, which corresponds to the threshold for viral culture^6,15^.

### Parameter estimation

For each model, we estimated simultaneously the vector of individual parameter *Ψ*_*i*_, which depends on both the parameters of the viral kinetic model (*R*_0_, δ, *p*, 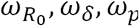, ω*δ*, ω*p*) and the parameters of the transmission model (*β*, ω_*β*_). The model providing the lowest BIC was retained. All parameters were estimated by computing the maximum-likelihood estimator using the stochastic approximation expectation-maximization (SAEM) algorithm implemented in Monolix Software 2020R1 (http://www.lixoft.eu/)^27–29^.

### Simulations settings

We provided prediction intervals for viral load and transmission probability over time, depending on the nature of contact, namely household (*h* = 1) or not (*h* = 0). In this purpose, we performed simulations, sampling *M* = 1000 individual vectors of parameters *Ψ*_*m*_ from the population distribution. Then, we derived the predicted viral load 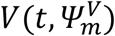 and the predicted transmission probability at all times according to the type of contact *P*_*h*_(*t, Ψ*_*m*_).

We then calculated the median viral load, 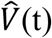 and the median transmission probability 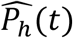 over the *M* simulated individuals, as well as the first and third empirical quantiles to provide prediction intervals.

All simulations were performed using the Simulx package on R.3.6.0.

### Calculating the average probability of transmission

Using our model, we also aimed to visualize the impact of a therapeutic intervention or a virus mutation on the probability of transmission, first at the individual level at each time, and second at the population level as an average transmission probability during the 13 days period where contacts occurred (from day −5 to day 7 post-symptoms onset). In this purpose, we defined several scenarios of simulation by modifying the corresponding parameters in the viral dynamic model. First, we increased the viral production parameter, *p*, by a factor of 2, 4, 8,16, 32, 64 and 100 corresponding to observed increases of 1-7 *C*_*t*_ value for different variants^2,10,11^. Second, we decreased the production parameters *p* by a factor of 2, 4, 8 and 16 as well^4^ to emulate the impact of vaccination^4,13^. Simulations were performed following same procedure as above. Then, for each emulated scenario, we computed the average probability of transmission as the empirical mean of transmission probabilities over the 13 days period where contacts occurred, weighted by the proportion of contacts at each day *p*_*t,h*_ (supplementary figure 2):

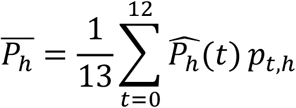

We computed the resulting increase or decrease of the probability of transmission compared to the baseline scenario.

## Supplementary Figures

**Supplementary Figure 1:**
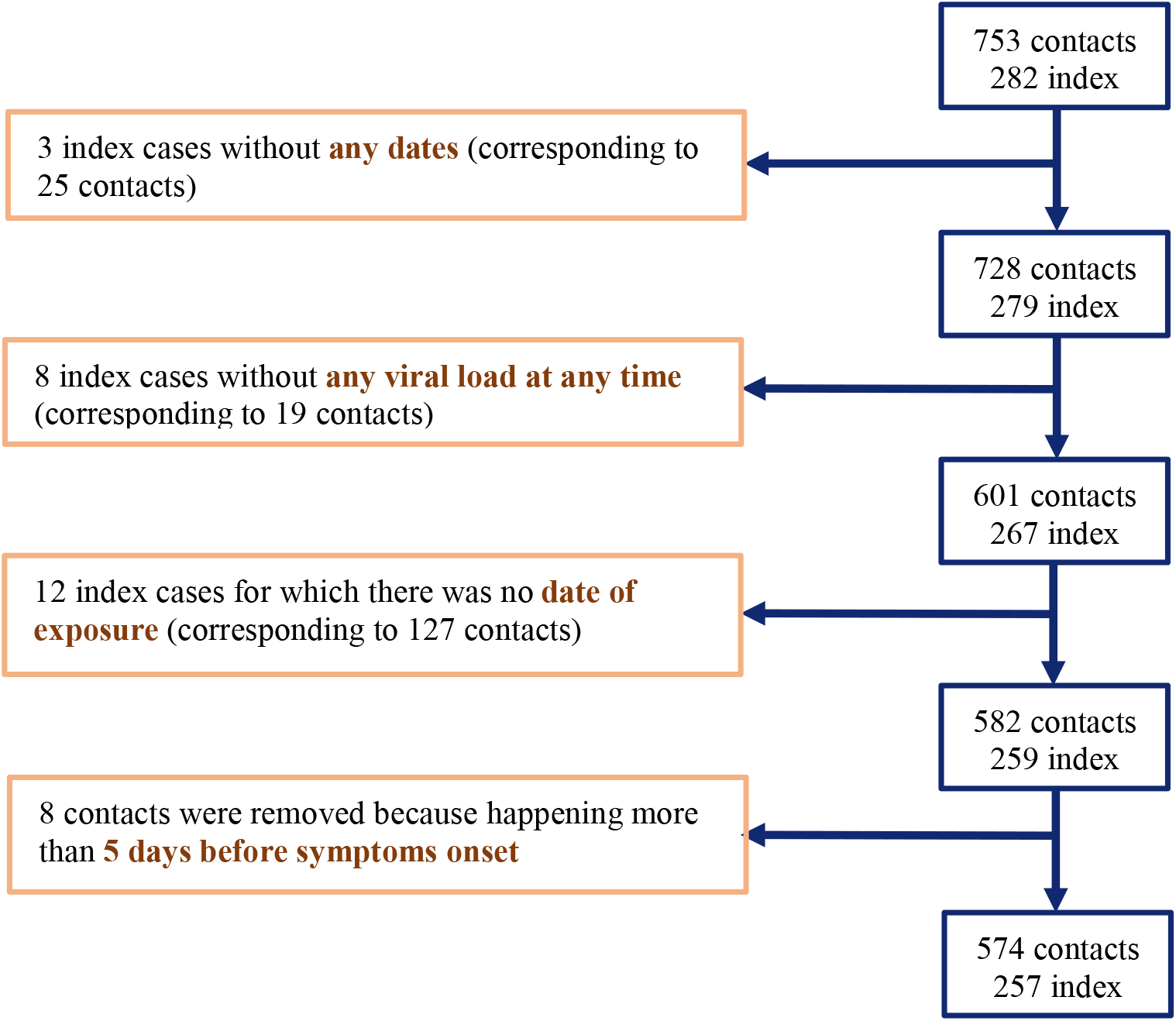
Flow chart of data selection.

**Supplementary Figure 2:**
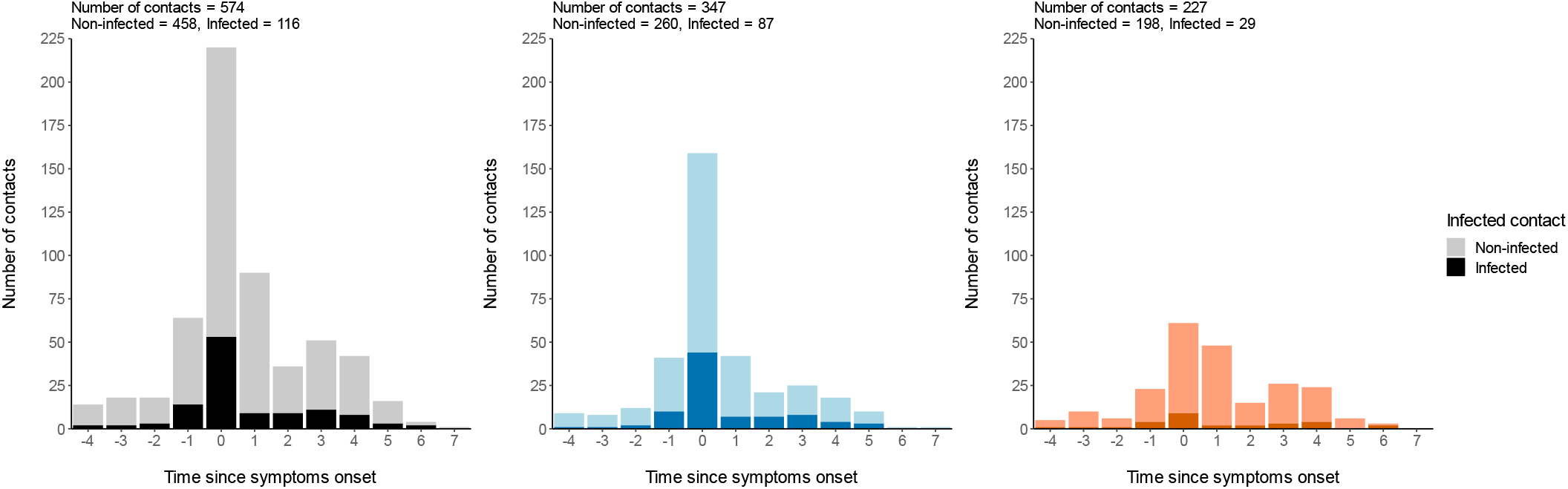
Distribution of contacts based on symptoms onset. All contact type (Left), Household contacts (Middle), Non-household contacts (Right). Contacts resulting in infection are represented as a darker shade.

**Supplementary Figure 3:**
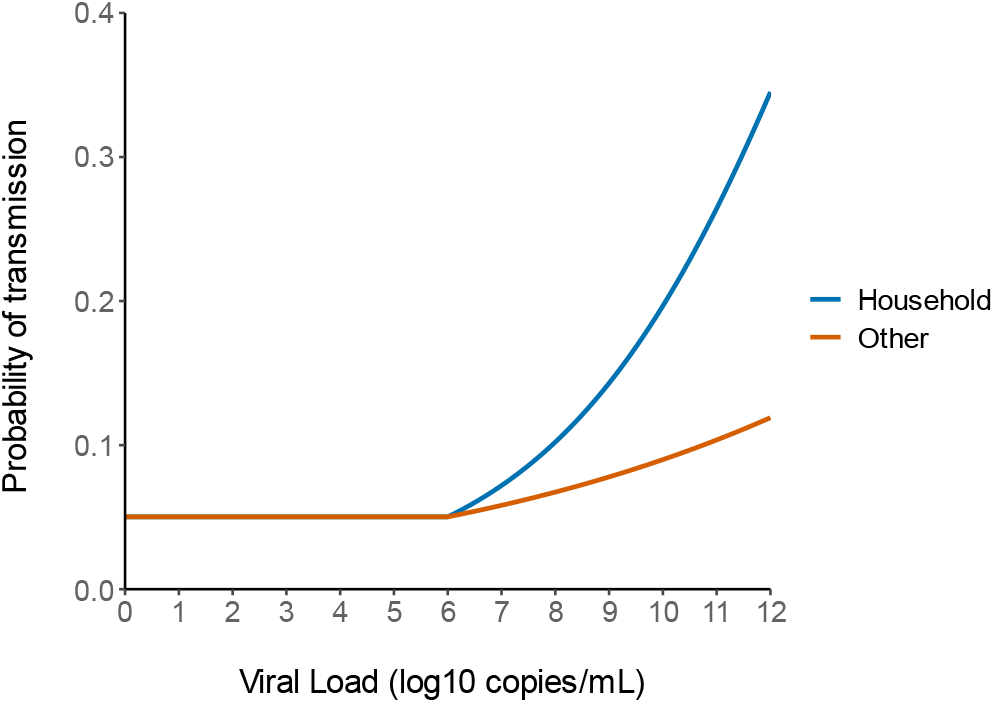
Predicted probability of transmission as a function of log-viral load using the population parameters of model M2.

## Supplementary Tables

**Supplementary Table S1:** Parameters estimates of the three candidate models. R_0_, basic reproductive number; δ, loss rate of infected cells; p, rate of viral production; γ_1_ represents the effect of household contacts on the transmission probability; γ_0_ represents the effect of non-household contacts on the transmission probability. M4 is a model where the transmission probability does not depend on the viral load. M5 is a model where the transmission probability depends on the maximum observed viral load for the index case.

| Parameter estimates (RSE %) |  |  |  |  |
| --- | --- | --- | --- | --- |
| No effect of viral load with additive variability (M4) |  |  | Logit-Linear with MaxVL (M5) |  |
|  | Fixed effect | Random effect SD | Fixed effect | Random effect SD |
| $R_0$ | 14.5 (53) | 0.327 (72) | 32.5 (51) | 0.42 (127) |
| $\delta \text{ (d}^{-1}\text{)}$ | 0.843 (2.81) | 0.05 (46) | 0.857 (16) | 0.06 (200) |
| $p \text{ (cells}^{-1}\text{.d}^{-1}\text{)}$ | $3.3 \times 10^5$ (136) | 2.53 (16) | $1.43 \times 10^6$ (189) | 2.61 (12) |
| $\gamma_1$ | 1.6 (12) | 1.19 (27.2) | 0.358 (35) | 1.4 (31.8) |
| $\gamma_2$ | 0.663 (46) | | 0.124 (79) | |
| $BIC$ | 2494 | | 2500 | |

**Supplementary Table S2:** Impact of changes in viral production rate on the average transmission probability during a high-risk contact..

| Average probability of transmission %<br>(Difference with the predicted value) |  |  |  |  |  |  |  |  |  |  |  |  |
| --- | --- | --- | --- | --- | --- | --- | --- | --- | --- | --- | --- | --- |
| Logit-Linear (M2) |  |  |  |  |  |  | Log-linear Model (M3) |  |  |  |  |  |
| Increased viral production |  |  |  | Decreased viral production |  |  | Increased viral production |  |  | Decreased viral production |  |  |
|  | Household | Non-Household | Peak viral load (log<sub>10</sub> copies per mL) | Household | Non-Household | Peak viral load (log<sub>10</sub> copies per mL) | Household | Non-Household | Peak viral load (log<sub>10</sub> copies per mL) | Household | Non-Household | Peak viral load (log<sub>10</sub> copies per mL) |
| Observed value | 25.1 | 12.8 | - | 25.1 | 12.8 | - | 25.1 | 12.8 | - | 25.1 | 12.8 | - |
| Predicted value | 24.7 | 11 | 9.4 | 24.7 | 11 | 9.4 | 25.1 | 12.2 | 9.3 | 25.1 | 12.2 | 9.3 |
| Fold change in p |  |  |  |  |  |  |  |  |  |  |  |  |
| 2 | 26.8<br>(+8.4) | 11.9<br>(+7.2) | 9.7<br>(+3.2) | 22.7<br>(-8.4) | 10.3<br>(-6.9) | 9.1<br>(-3.2) | 28<br>(+11.5) | 13.4<br>(+9.5) | 9.6<br>(+3.2) | 22.3<br>(-11.1) | 11.2<br>(-8.8) | 9<br>(-3.2) |
| 4 | 28.9<br>(+16.8) | 12.7<br>(+14.8) | 10<br>(+6.4) | 20.6<br>(-16.6) | 9.6<br>(-13.5) | 8.8<br>(-6.4) | 31<br>(+23.4) | 14.7<br>(+19.7) | 9.9<br>(+6.5) | 19.7<br>(-21.3) | 10.2<br>(-16.7) | 8.7<br>(-6.5) |
| 8 | 30.9<br>(+25) | 13.6<br>(+22.5) | 10.3<br>(+9.6) | 18.6<br>(-24.8) | 8.9<br>(-19.6) | 8.5<br>(-9.6) | 34<br>(+35.5) | 16<br>(+30.5) | 10.2<br>(+2.1) | 17.4<br>(-30.8) | 9.4<br>(-23.7) | 8.4<br>(-9.7) |
| 16 | 32.9<br>(+33.2) | 14.5<br>(+30.4) | 10.6<br>(+12.8) | 16.7<br>(-32.6) | 8.3<br>(25.3) | 8.2<br>(-12.8) | 37.1<br>(+47.8) | 17.4<br>(+42.1) | 10.5<br>(+12.9) | 15.2<br>(-39.4) | 8.6<br>(-30) | 8.1<br>(-12.9) |
| 100 | 38.0<br>(+53.9) | 16.8<br>(+52.1) | 11.4<br>(+21.3) | 12.1<br>(-50) | 6.9<br>(-38) | 7.4<br>(-21.3) | 45.2<br>(+80) | 21.5<br>(+76) | 11.3<br>(+21.5) | 10.6<br>(-57.5) | 7<br>(-43) | 7.3<br>(-21.5) |

## Notes

### Competing Interest Statement

The authors have declared no competing interest.

### Funding Statement

The study has received financialsupport from the National Research Agency (ANR) through the ANR-Flashcall for COVID-19 (Grant ANR-20-COVI-0018) and the Bill and Melinda GatesFoundation under Grant Agreement INV-017335. The original trial was funded by a crowdfunding campaign YoMeCorono (https://www.yomecorono.com/), and Generalitat de Catalunyawith support for laboratory equipment fromFoundation Dormeur. We thank Samuel Alizon, Xavier Duvaland Xavier de Lamballerie for helpful discussions.

### Author Declarations

The data used in this study originate from an original clinical study. Details on the study can be found in : Transmission of COVID-19 in 282 clusters in Catalonia, Spain: a cohort study. The trial protocol and subsequent amendments were approved by the institutional review board at Hospital Germans Trias i Pujol and the Spanish Agency of Medicines and Medical Devices. All the participants provided written informed consent.

